# Designing and implementing state-level fertility preservation health insurance benefit mandates

**DOI:** 10.1101/2021.06.13.21258849

**Authors:** Ricardo E. Flores, Sara W. Yoeun, Omar Mesina, Bonnie N. Kaiser, Sara B. McMenamin, H. Irene Su

**Author notes:** Corresponding Author H. Irene Su, MD, MSCE, Moores Cancer Center, University of California, San Diego, 3855 Health Sciences Drive, Dept 0901, La Jolla, CA 92093-0901. **Capsule** Variation was found among the design and implementation of 11 state-level fertility preservation health insurance mandates. Best practices for stakeholders to consider when designing and implementing benefit mandates are described.

## Abstract

**Objective:** To describe the design and implementation of state-level fertility preservation (FP) health insurance benefit mandates and regulation and to provide stakeholders with guidance on best practices, gaps, and implementation needs.

**Design:** Legal mapping and implementation framework-guided analysis

**Setting:** U.S. states with state-level fertility preservation health insurance benefit mandates

**Patient(s):** Individuals at risk of iatrogenic infertility

**Intervention(s):** State laws mandating health insurance benefit coverage for fertility preservation services.

**Main Outcome Measure(s):** Design features of FP mandated benefit legislation; implementation process

**Result(s):** Between June, 2017 and March, 2021, 11 states passed FP benefit mandate laws. On average, states took 223 days to implement their mandates from the start of the laws’ enactment dates to their corresponding effective dates, and a majority issued regulatory guidance after the law was in effect. Significant variation was observed in which FP services were specified for inclusion or exclusion in the laws and/or regulator guidance. Federal policies impacted state level implementation, with the ACA and HIPAA guiding design of fertility preservation benefits. In addition, a majority of states referenced medical society clinical practice guidelines in the design of FP mandated benefits.

**Conclusions:** Our policy scan documented significant variation in the design and implementation of health insurance benefit mandates for FP services. Future considerations for policy development include specificity and flexibility of benefit design, reference to external clinical practice guidelines to drive benefit coverage, inclusion of Medicaid populations in required coverage, and consideration of interaction with relevant state and federal policies. In addition, key considerations for implementation include the sufficient length of time for the implementation period, regulator guidance issued prior to the law going into effect, and explicit allocation of resources for the implementation process.

## Introduction

Nearly 70,000 young people aged 0 to 39 are diagnosed with cancer annually.^1,2^ These pediatric, adolescent, and young adult cancer patients can experience higher risks of infertility due to cancer treatments. Infertility is a critical area of concern for many young cancer survivors, but risks can be reduced by the evidence-based practice of fertility preservation (FP) care. However, the expense of standard FP services can be prohibitive and are a common barrier to care. Thus, state-level health insurance benefit mandates for FP care have been introduced in recent years to increase access and utilization to these services.

Since 2017, 11 states (California [CA], Colorado [CO], Connecticut [CT], Delaware [DE], Illinois [IL], Maryland [MD], New Hampshire [NH], New Jersey [NJ], New York [NY], Rhode Island [RI],and Utah [UT]) have passed health insurance benefit mandates for coverage of FP services, and more states have legislation under consideration.^3^ This legislation requires health insurers to include coverage for FP services in specified types of health insurance plans. After passage, implementation of benefit mandates require multiple steps. Benefit mandates are interpreted by state insurance regulatory agencies, which may issue guidance for implementation by insurers subject to the law. Insurers then design benefits that comply with the law and insurance regulator guidance and may inform contracting healthcare providers and patients of newly covered services. Finally, patients and clinics (i.e., healthcare providers and representatives) communicate with the insurers to determine how to access the benefits and to understand the terms of coverage for services.

To date, patients, advocates, healthcare providers, medical societies and policymakers have championed these laws, but the effectiveness of this health policy intervention in increasing access to FP services is unknown.^4-11^ Healthcare providers in states with benefit mandate laws have experienced significant variation in and confusion around accessing insurance coverage for patients. This suggests problems with either the intervention (i.e., benefit mandate law) and/or implementation at the state-, insurer-, clinic-, or patient-level. Key unanswered questions include how heterogeneous are benefit mandate laws and their regulation, and how might this variation impact implementation, access, and utilization.

The objectives of this study are to systematically characterize the variation in state-level FP health insurance benefit mandates and regulation using legal mapping^12^ and implementation science methods^13^ and to provide stakeholders (e.g., patients, healthcare providers, insurers, and policymakers) with guidance on best practices, gaps, and implementation needs. In this context we took a novel approach of conducting a policy scan—a component of legal mapping—to identify the heterogeneity and consistencies in state-level benefit mandate laws and regulatory guidance. We then conducted a systematic, implementation framework-guided evaluation focused on how the benefit mandates and regulatory guidance interact with federal laws, medical societies, and insurers to influence implementation.

## Methods

### Development of dataset

Collection of the legislative and regulatory policies used in this policy scan was conducted in two stages. First, a list of states who have passed FP mandates was identified through the Alliance for FP.^3^ This identified 11 states that had passed legislation as of March 22, 2021. For each state on this list, we downloaded final versions of legislative text from the state legislature’s website in PDF format.

Additional searches were conducted on insurance regulators’ websites in each of the 11 states to identify FP legislation-specific guidance generated by insurance regulators to insurers. Terms used in this search included: “fertility preservation,” “all-plan letter,” “bulletin,” and “insurance check list.” To validate the search results, email and/or telephone inquiries were made to each regulator to confirm that the regulator documents that were identified from the web-search were the only guidance from the regulators to the insurers related to the implementation of the FP benefit mandate. Both the legislative text (n=11) and regulator documents (n=10) were uploaded to MAXQDA 2020.^14^ This research evaluated public programs and did not involve human subjects and thus was exempt from IRB review.

### Code development and coding

The authors developed a list of deductive (*a priori*) codes based on the Exploration, Preparation, Implementation, Sustainment (EPIS) implementation science framework.^13^ Implementation science, which seeks to integrate interventions into clinical settings to improve patient outcomes, is guided by frameworks including EPIS that serve as a map to conduct a systematic inquiry of key domains that impact implementation. The EPIS framework was selected because it explicitly identifies the importance of the innovation (FP health insurance benefit mandate law), outer context (insurance regulators, federal law, medical societies), inner context (insurers), and bi-directional bridging factors (e.g., insurance regulator guidance, clinical practice guidelines) within and between these contexts that impact implementation of the law. An initial review of the legislative and regulator documents led to further development of inductive codes (i.e., themes that arose from the laws and regulator guidance).

Using the preliminary code book, four authors (RF, SY, SM, HIS) coded the legislative and regulator documents for two selected states. Coding discrepancies were reviewed, and codes were revised into a final codebook. Two coders (RF, SY) then independently coded all of the documents using MAXQDA 2020.^14^ Intercoder agreement was above 80%, and all coding discrepancies were resolved through discussion.

### Data analysis

Data were summarized by theme (i.e., code), with structured comparison of each theme across states.^15^ In legislative and regulator documents, references were made to medical society guidelines (ACOG, ASRM, ASCO) and essential health benefits (EHB)-benchmark plans. Guidelines from ACOG, ASRM and ASCO were subsequently located via web searches, and content was reviewed as needed to clarify legislative and/or regulatory language.^6,7,9^ Clarifying text was incorporated into theme summaries. In addition, the Centers for Medicare and Medicaid Services website was also searched to provide better context for the mention of EHBs in the legislative and/or regulatory language ^16^

## Results

Between June, 2017 and March, 2021, 11 states passed FP benefit mandate laws (**Table 1**). Legislative text were obtained for all 11 states. Insurance regulator guidance were obtained for all states except NJ and UT. There were two regulator documents for NY. Overall, 21 documents were included in the analysis.

**Table 1:** Summary of fertility preservation benefit mandate enactment, insurance regulator communication, and insurance market segments impacted by state.

|  | Benefit mandate enactment date | Benefit mandate effective date | Regulator communication date | Days between enactment date and |  | Commercial market plans |  |  | Public plans |
| --- | --- | --- | --- | --- | --- | --- | --- | --- | --- |
|  |  |  |  | Effective date | Regulator communication date | Individual | Small Group<sup>1</sup> | Large Group | Medicaid |
| **CA** | 10/12/2019 | 10/12/2019<sup>3</sup> | 01/15/2020 | 0 | 95 | ? | ? | ? |  |
| **CO** | 04/01/2020 | 01/01/2022 | 07/30/2020 | 640 | 121 | ? | ? | ? |  |
| **CT<sup>4</sup>** | 06/20/2017 | 01/01/2018 | 03/19/2019 | 195 | 637 |  | ? | ? |  |
| **DE** | 06/30/2018 | 06/30/2018 | 10/09/2018 | 0 | 101 | ? |  | ? |  |
| **IL** | 08/27/2018 | 01/01/2019 | 05/01/2020 | 127 | 613 | ? | ? | ? | ?<sup>5</sup> |
| **MD** | 05/15/2018 | 01/01/2019 | 01/01/2019 | 23 | 231 |  |  | ? |  |
| **NH** | 08/05/2018 | 01/01/2020 | 04/13/2020 | 153 | 617 |  | ? | ? |  |
| **NJ** | 01/13/2020 | 04/12/2020 | N/A | 90 | N/A |  | ? | ? |  |
| **NY<sup>4</sup>** | 01/13/2019 | 01/01/2020 | 12/3/2019 | 264 | 324 | ? | ? | ? |  |
| **RI** | 07/05/2017 | 07/5/2018 | 12/08/2017 | 365 | 156 | ? | ? | ? |  |
| **UT<sup>6</sup>** | 5/16/2021 | 1/1/2023 | N/A | 594 | N/A |  |  |  | ? |

? = Included
<sup>1</sup>50 or fewer full-time equivalent employees.
<sup>2</sup>Health plans in place before March 23, 2010, when the Affordable Care Act was signed into law are not allowed to reduce their benefit coverage or they lose their grandfathered status.
<sup>3</sup>Policy applies retroactively to plans from January 1, 1990, onward
<sup>4</sup>Coverage limited to individuals who previously maintained coverage under the policy for at least 12 months
<sup>5</sup>Legislation does not specify Medicaid coverage, but regulator guidance does
<sup>6</sup>Pending Centers for Medicare & Medicaid Services approval

### Policy scan of laws and insurance regulator guidance

The enactment date (i.e., date the law was passed), effective date (i.e., date the law took effect), regulator communication date, and insurance market segments impacted by FP benefit mandates are presented for each state in **Table 1**. On average, states had 223 days to implement their FP benefit mandates (range 0 to 640 days). Among state insurance regulators that issued guidance, regulators took an average of 322 days to provide guidance related to implementation of FP benefit mandates (range 95 to 617 days). Four of the nine regulators issued communication prior to the effective date of the mandate. Ten out of 11 laws apply to commercial large group (i.e., 51 or more employees) plans with variation in requirements for individual and small group plans. IL is the only state law with coverage applicable to both commercial and state Medicaid plans, while UT’s law only applies to Medicaid benefits.

#### How is iatrogenic infertility defined?

Laws in 10 states (excluding UT) and/or regulator guidance (excluding NJ) include a definition for iatrogenic infertility, specifying that iatrogenic infertility can stem from treatments that directly or *indirectly* cause infertility. A majority of states (excluding MD and DE) cited not only cancer treatments (i.e., chemotherapy, radiation, surgery) but also “*other”* treatments that could cause infertility. Five states (CA, MD, NJ, NY, RI) referred to medical societies to set standards for medical treatments that can cause infertility.

#### What FP services are covered?

Standard of care FP procedures such as embryo cryopreservation require medical evaluation, medications, ultrasound and laboratory monitoring, oocyte retrieval, cryopreservation, and storage. There was significant variation in which of these services were specified for inclusion or exclusion in the laws and/or regulator guidance (**Table 2**). DE law included the broadest scope of specified covered services. In contrast, CA policy did not define coverage for any specific services. Where oocyte and sperm freezing were specified for seven of the same states, embryo freezing was specified in only three states (DE, NH, UT). Four states specified coverage for storage, while three states specified exclusion of storage. Four states (CO, CT, MD, RI) set limits on total costs or number of cycles. All states—excluding CT—referred to clinical society guidelines to set what is considered standard of care FP procedures, allowing for coverage of future standard of care procedures that may develop over time.

**Table 2:**
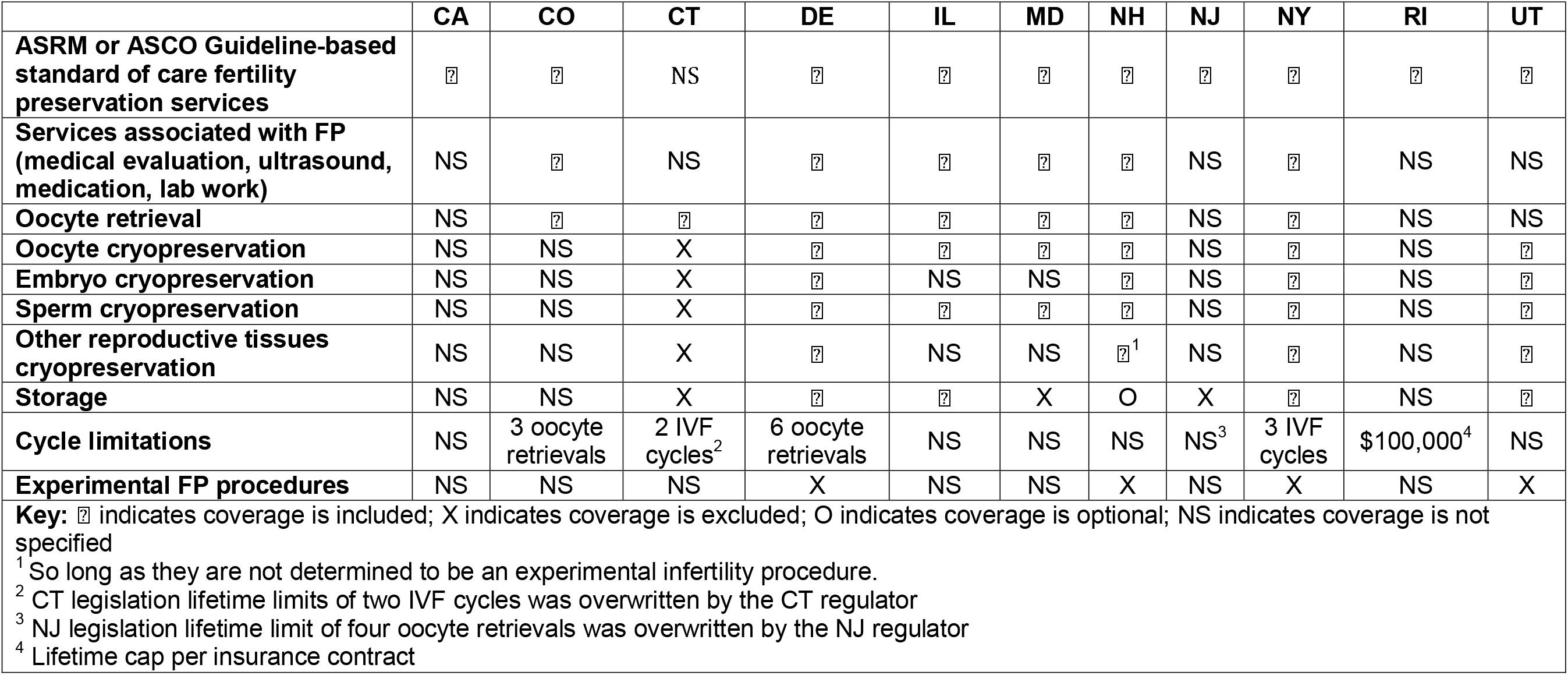
Summary of variation in health insurance benefit mandate coverage specifics for fertility preservation for iatrogenic infertility by state.

#### Additional limitations and prevention of restrictions

Four states (CO, CT, DE, MD) allow religious employers to request exemptions for coverage based on their religious beliefs. CT expands on this to include individuals with religious or moral beliefs that are contrary to diagnosis and treatment of infertility including FP and specified that they are eligible for insurance policies that do not include these benefits.

Most states did not specify age restrictions, with the exceptions of DE law (oocyte retrievals before age 45), IL regulator guidance (age 14-45), and NY law (age 21-44). While NY legislation included age limitations, the guidance that followed from the state insurance regulator indicated that age restrictions are not permitted. Three states (IL, NJ, NY) state that coverage cannot be restricted based on life expectancy or predicted disability.

#### How is cost-sharing addressed?

Cost-sharing (i.e., out-of-pocket costs incurred by the patient when they use FP services covered by their health insurance plan), was described by six states’ (CO, CT, DE, NH, NJ, NY) legislation or regulator guidance. Five states (CO, DE, NH, NJ, NY) included language on parity to prevent insurers from charging more (via deductibles, copayments, and/or coinsurance) or putting more restrictions (e.g., benefit maximums and waiting periods) on FP benefits that are not in place for other medical services. In contrast, CT regulator guidance allowed insurers to apply plan level cost-sharing mechanisms, coinsurance of up to 50%, and prior authorization. For both CT and NY, cost-sharing is subject to regulator oversight.

### Implementation of benefit mandates

The EPIS framework was used to describe key domains that impact implementation of benefit mandates. Within the framework, benefit mandate legislation is implemented in the inner context (i.e., the health insurer) and is influenced by the outer context (i.e., related state and federal health insurance policies, medical societies) and bridging factors (i.e., regulator interpretation and communication) between the inner and outer contexts. Themes within these domains are discussed below, with no inner context themes identified.

#### Outer context

Federal laws such as the Affordable Care Act (ACA) and Health Insurance Portability and Accountability Act (HIPAA) are components of the outer context that impact state-level benefit mandates. The ACA established a mechanism for determining essential health benefits (EHBs) required to be covered in non-grandfathered individual and small group market plans.^17^ It is difficult to pass benefit mandate legislation if it is interpreted to exceed EHBs, because an ACA provision requires states to defray costs of providing benefits if the passed benefit mandate exceeded the state-defined EHB benchmark plan. Thus, CO and CA laws made arguments that FP mandates do not exceed EHBs because they are an additional service covered under an existing pregnancy benefit (CO) or are already covered as a “basic healthcare service” (CA). Should the federal government determine that FP services exceed EHBs and that the state must provide the cost of coverage for FP services, states including CO and IL included language that would render the mandates inoperable.

Two additional federal laws were referenced in either legislation or regulator guidance. CT insurance guidance clarified that age limits are discriminatory if applied to services that are clinically effective at non-included ages, which is not permissible under section 1557 of the ACA. Also in CT and NJ insurance guidance, lifetime limits on FP benefits were determined to function as an “impermissible preexisting condition exclusion” under HIPAA.

#### Bridging factors

Bridging factors centered on communication across levels. Explicit reference to clinical practice guidelines in the legislation or regulator guidance bridges the gap between the medical societies in the outer context and the insurers in the inner context. Laws in 10 states referred to ACOG, ASRM, and/or ASCO guidelines to set procedures and services that are consistent with standard of care for FP. In addition, laws in five states (CA, MD, NJ, NY, RI) referred to clinical societies to set standards for medical treatments that can cause infertility. Third, ASRM standards were referenced to guide insurers until the NH regulator could “adopt necessary rules” to administer the law. Finally, the NY law designated regulators to design regulations in accordance with standards and guidelines adopted by ACOG and ASRM.

Regulator guidance also served as a bridging factor between regulators in the outer context and insurers in the inner context. Several themes within regulator guidance demonstrated how state insurance regulators influence implementation. Guidance explained coverage beyond the legislative language. NH, NY, IL, and CT regulator guidance all added coverage details beyond the legislation. For example, NY regulator guidance clarified that FP services are a separate benefit that did not count toward the 3-cycle limit on IVF benefits for infertility, as well as that storage is a covered benefit. Second, discordance between the mandate benefit legislation and federal regulations were noted in multiple regulator documents. For example, CT guidance instructed insurers on the discrepancy and then instructed insurers to remove age limits on FP benefits and the requirement that enrollees disclose previous services received in their lifetime. Finally, guidance specified informing insured members on the benefit and communicating with regulators for compliance. In California, the all-plan letter from the insurance regulator instructed insurers to review and modify a range of consumer-facing documents (i.e., Explanation of Benefits, Summary of Benefits and Coverage, Schedules of Benefits, Infertility Riders, Subscriber Agreements, and disclosure forms) to ensure compliance with the law. Subsequently the regulator requires insurers to communicate their plans for ensuring compliance. On additional insurer to patient guidance, CT law detailed that insurance policies that do not include FP services shall provide written notice of “not less than ten-point type, in the policy, application and sales brochure for such policy” to insured and prospective insured patients.

## Discussion

Significant prior and ongoing efforts to support health insurance coverage of FP services have resulted in state-level insurance benefit mandates with heterogeneity that may influence implementation, and ultimately, patients’ access to care. We characterized variation in the benefit mandates, documented how benefit mandates are influenced by external factors, and in turn, how regulator communication and clinical practice guidelines can bridge the gap between the external policy context and implementation by health insurers. The legal mapping and implementation science-informed approach yielded a systematic, novel evaluation of these health policies to update stakeholders on potential determinants of successful implementation and current gaps.

### Benefit Mandate Design

Several best practices for FP benefit mandate design emerged from this policy scan. First, all 11 mandates reviewed in this study were “mandates to cover” rather than “mandates to offer.” A “mandate to *cover*,” in which all benefit designs from an insurer have to include the benefit, is distinct from a “mandate to *offer*,” in which only one benefit design would need to include a FP benefit, leaving the vast majority of patients without FP coverage.

Second, legislation should include language that establishes FP benefits at parity with other covered health care services. This results in treating individuals who need FP services fairly, by preventing insurers from charging more or putting more restrictions of FP benefits compared to other medical care.

A third best practice is that FP legislative language needs to be specific yet flexible such that the scope of services to be covered is not up to interpretation by health insurers but is also responsive to the needs of each individual. Language that is not specific can lead to heterogeneity which leads to a patchwork of benefits and potentially, a more restricted scope of coverage defined by insurers which may not include important benefits such as IVF and storage of cryopreserved material. Language that specifies flexibility on the number of FP cycles would ensure that coverage can be robust enough such that it is meaningful; however, this was not observed in most legislation to date. More systematic evaluation comparing benefit designs and patient-level benefit utilization among states by specification of scope of coverage are needed to inform the most effective approach.

Finally, FP coverage needs to be offered to the populations that need it the most. FP is currently a benefit that is only available to those with the means to pay the high costs of the services. Only two of the 11 states that mandate coverage for FP services include the lowest income populations (i.e., Medicaid beneficiaries) under their mandate. To ensure health equity across populations, the inclusion of the Medicaid population in FP benefit mandates is essential.

### Implementation

We observed significant variation (of up to two years) in the implementation period between the laws’ enactment dates and their corresponding effective dates. This time is crucial for health insurers to make modifications to their standard benefit plans and communicate changes to their contracting providers and enrollees. In addition, only one state (CO) had legislation that appropriated funds for the implementation process. This legislation allocated state funds to pay for department of regulatory agencies staff salary—equal to 0.1 FTE—to implement the policy as laid out in the law.

### Outer context constraints and facilitators

Outer context constraints identified include state and federal health insurance policy. The process by which each state selects an EHB-benchmark plan provides an opportunity for FP benefits and infertility benefits to be defined as an EHB. This is important in that mandated benefits that are perceived to exceed the state-defined EHBs post an additional price tag to the state. Of the 11 states that were included in this study, eight included coverage for infertility services in their 2017 EHB-benchmark plan (CO, CT, IL, MD, NH, NJ, NY, RI).^16^ While no state currently includes explicit coverage for FP in their benchmark plan, it is possible to argue that a FP mandate does not exceed EHBs as it is a clarification of an existing covered infertility benefit. Crafting an argument for how proposed FP benefit mandates do not exceed EHBs is an essential strategy for such legislation to have political viability. In addition, leveraging federal ACA and HIPAA antidiscrimination laws, advocates can push back against language that requires lifetime or age limitations.

### Bridging factors as facilitators

Insurance regulators are a key player in benefit mandate implementation. The EPIS framework’s bridging construct enabled characterization of how regulators bridged mandates with state and federal policies and health insurance plans. Our policy scan showed that regulator guidance varied in scope and frequently highlighted how state legislation conflicted with federal law (e.g., age restrictions, and lifetime limitations). More than half of the regulators issued guidance related to implementation of the FP mandate after the mandate went into effect. To have a more significant impact on the implementation process, regulator guidance should be issued prior to the mandate effectiveness date. Regulator communication represents an opportunity to ensure compliance with existing laws as well as specify and update scope of coverage. More data are needed to determine how insurance regulators shape their guidance documents.

Through clinical practice guidelines, medical societies that set care standards based on clinical expertise and best evidence can have significant impact on health policy interventions. National clinical practice guidelines enable re-categorization of experimental to standard of care procedures over time (e.g., ovarian tissue freezing) that may then be included in benefits for standard of care FP services without each state specifying a new service. With little language specifying the types and number of services to be covered as standard of care within present day laws and regulator guidance, there is an opportunity for medical societies—through the revision of current guidelines—to expand coverage to meet the breadth of needs and indications in patients. As well, many laws refer to guidelines to define the at-risk population, and guidelines do not currently specify which treatments may cause infertility, leaving room for broad or narrow interpretations by insurers. If clinical practice guidelines do not better define at-risk populations, then laws should not refer to them.

Several strengths of this work include use of implementation science and legal mapping methodology to systematically evaluate laws pertinent to fertility care, inclusion of insurance regulator guidance to complement laws, and an update of current FP benefit mandates with a focus on variations among states. The limitation is that the study did not evaluate insurer-, clinic- and patient-level data on benefit mandate implementation, which is needed to identify determinants, moderators, and mediators of patient utilization of FP benefits within benefit mandates and insurance regulator guidance.

## Conclusions

Our policy scan documented significant variation in the design and implementation of health insurance benefit mandates for FP services across 11 states with enacted laws. As future states consider enacting FP mandated benefits, key considerations for policy development include mandates to provide coverage, specificity and flexibility of benefit design, limitations in referencing clinical practice guidelines, inclusion of Medicaid populations in required coverage, and consideration of interaction with relevant state and federal policies. In addition, key considerations for implementation include the sufficient length of time for the implementation period, regulator guidance issued prior to the law going into effect, and explicit allocation of resources for the implementation process.

## Data Availability

The data that support the findings of this study are openly available via state legislature and insurance regulator websites

## Acknowledgements

This work was funded by UC San Diego Moores Cancer Center, Specialized Cancer Center Support Grant NIH/NCI P30CA023100 and UC San Diego Academic Senate pilot grant.

## Notes

### Competing Interest Statement

HIS received speaking honorarium from Ferring Pharmaceutical

### Author Declarations

Research evaluated public programs and did not involve human subjects and thus was exempt from IRB review.

## References

1. Surveillance, Epidemiology, and End Results (SEER) Program. SEER*Stat Database: Incidence - SEER 18 Regs Research Data + Hurricane Katrina Impacted Louisiana Cases, Nov 2016 Sub (1973-2014 varying) - Linked To County Attributes - Total U.S., 1969-2015 Counties, National Cancer Institute, DCCPS, Surveillance Research Program, released April 2017, based on the November 2016 submission. www.seer.cancer.gov/seerstat

2. American Cancer Society. Cancer Treatment and Survivorship: Facts and Figures 2020. 2020.

3. Alliance for Fertility Preservation. State Laws & Legislation. Accessed March 2021, https://www.allianceforfertilitypreservation.org/advocacy/state-legislation

4. Patel P, Kohn TP, Cohen J, Shiff B, Kohn J, Ramasamy R. Evaluation of Reported Fertility Preservation Counseling Before Chemotherapy Using the Quality Oncology Practice Initiative Survey. JAMA Netw Open. Jul 1 2020;3(7):e2010806. doi:10.1001/jamanetworkopen.2020.10806

5. California Health Benefits Review Program. Analysis of Senate Bill (SB) 600: Fertility Preservation. A report to the 2019-2020 California State Legislature. April, 2019 2019;

6. Oktay K, Harvey BE, Partridge AH, et al. Fertility Preservation in Patients With Cancer: ASCO Clinical Practice Guideline Update. J Clin Oncol. Jul 1 2018;36(19):1994–2001. doi:10.1200/JCO.2018.78.1914

7. Practice Committee of the American Society for Reproductive Medicine. Electronic address aao. Fertility preservation in patients undergoing gonadotoxic therapy or gonadectomy: a committee opinion. Fertil Steril. Dec 2019;112(6):1022–1033. doi:10.1016/j.fertnstert.2019.09.013

8. National Comprehensive Cancer Network. NCCN Clinical Practice Guidelines in Oncology: Adolescent and Young Adult (AYA) Oncology, Version 1.2021-September 10, 2020. Accessed June 2, 2021, https://www.nccn.org/professionals/physician_gls/default.aspx#age

9. American Congress of Obstetrics and Gynecology. ACOG Committee Opinion No. 747: Gynecologic Issues in Children and Adolescent Cancer Patients and Survivors. Obstet Gynecol. Aug 2018;132(2):e67–e77. doi:10.1097/AOG.0000000000002763

10. Sax MR, Pavlovic Z, DeCherney AH. Inconsistent Mandated Access to Fertility Preservation: A Review of Relevant State Legislation. Obstet Gynecol. Apr 2020;135(4):848–851. doi:10.1097/AOG.0000000000003758

11. Kawwass JF, Penzias AS, Adashi EY. Fertility-a human right worthy of mandated insurance coverage: the evolution, limitations, and future of access to care. Fertil Steril. Jan 2021;115(1):29–42. doi:10.1016/j.fertnstert.2020.09.155

12. Burris S. How to write a legal mapping paper. Temple University Legal Studies Research Paper. 2020;(2018–10)

13. Aarons GA, Hurlburt M, Horwitz SM. Advancing a Conceptual Model of Evidence-Based Practice Implementation in Public Service Sectors. Administration and Policy in Mental Health and Mental Health Services Research. 2011/01/01 2011;38(1):4–23. doi:10.1007/s10488-010-0327-7

14. MAXQDA2020. 2020.

15. Ulin PR RE, Tolley EE. Qualitative methods in public health: a field guide for applied research. 1st ed. San Francisco: Jossey-Bass 2005.

16. Centers for Medicare and Medicaid Services. Information on Essential Health Benefits (EHB) Benchmark Plans. https://www.cms.gov/CCIIO/Resources/Data-Resources/ehb

17. Bagley N, Levy H. Essential health benefits and the Affordable Care Act: Law and process. Journal of health politics, policy and law. 2014;39(2):441–465.

